# A Phase 2, randomized, double-blind, placebo-controlled multi-center trial sub-study for the clinical effects of paridiprubart treatment in hospitalized critically ill patients with COVID-19 ARDS

**DOI:** 10.1101/2023.09.21.23295853

**Authors:** Blair Gordon, Fiona Allum, Michael Brooks, Nishani Rajakulendran, Emmanouil Rampakakis, John Sampalis, EB05 Study Investigators

## Abstract

**Background:** Coronavirus disease 2019 (COVID-19) mortality is predominantly due to acute respiratory distress syndrome (ARDS). There are currently limited treatment options for ARDS, a life-threatening condition with different etiologies, secondary to inflammation-induced lung injury. Paridiprubart is a monoclonal antibody that inhibits Toll-like Receptor 4 (TLR4), a key player in ARDS pathophysiology.

**Methods:** This was a prespecified sub-study of a randomized, double-blind, placebo-controlled, Phase 2 trial evaluating the efficacy and safety of paridiprubart in COVID-19 patients with ARDS receiving invasive mechanical ventilation and additional organ support. Efficacy outcomes were 28- and 60-day all-cause mortality, and improvement in COVID-19 severity and ventilation-free days at 28-days post-treatment.

**Results:** Thirteen (13) and twenty (20) patients received paridiprubart and placebo, respectively. The groups were comparable for demographics and baseline parameters, except for higher kidney failure incidence and use of immune modulators and antivirals, and lower corticosteroids use in the paridiprubart group. Mortality at 28-days post-treatment was 7.7% (1/13) in the paridiprubart group versus 40.0% (8/20) for placebo (OR=0.125; 95% CI, 0.013-1.160; P=0.067; P[bootstrap]=0.011). 60-day mortality was 23.1% (3/13) in paridiprubart-treated patients and 45.0% (9/20) in placebo patients (OR=0.367; 95% CI, 0.077-1.749; P=0.208; P[bootstrap]=0.162). Mean survival time was 55.78 days for paridiprubart recipients compared to 41.44 days for placebo patients (HR=0.386; 95% CI, 0.077-1.436; P=0.156; P[bootstrap]=0.083). Although not statistically significant, results for other efficacy measures favored paridiprubart. Incidence of adverse events was similar in both groups.

**Conclusions:** In COVID-19 patients with ARDS requiring invasive ventilation and organ support, paridiprubart was efficacious in preventing mortality and improving clinical outcomes, with no safety concerns.

## INTRODUCTION

Coronavirus disease 2019 (COVID-19) has caused more than six million deaths globally, with most deaths being attributed to acute respiratory distress syndrome (ARDS)^1^, a condition caused by injury to the alveolar structure and surrounding microvasculature. Cases of severe COVID-19 have been associated with dysregulated host immune responses^2,3^. Specifically, severe acute respiratory distress syndrome coronavirus 2 (SARS-CoV-2) infection leads to the overactivation of the innate immune system’s Toll-like Receptor 4 (TLR4) by pathogen-associated molecular patterns (PAMPs) and host-derived damage associated molecular patterns (DAMPs)^4^. TLR4 overactivation subsequently triggers the excessive release of pro-inflammatory cytokines (cytokine release syndrome; CRS), leading to acute lung injury (ALI) and ARDS^5,6^.

There are currently two vaccines approved for the prevention of COVID-19 infections. However, the real-world effectiveness of the vaccines remains suboptimal with important rates of breakthrough cases, despite very high uptake of multiple vaccinations. Furthermore, the efficacy of vaccines against new COVID-19 strains remains unproven. Antiviral agents used for symptomatic patients in a non-hospitalized setting can be effective in reducing viral load and controlling the severity of the symptoms, but only when administered early in the infection before any respiratory distress has occurred. Once respiratory distress has begun, the aim of treatment should be modulation of the immune response and reversal of the lung damage. Despite the availability of the antiviral remdesivir^7,8^ and anti-inflammatory/immunomodulators, including dexamethasone^9,10^, anti-interleukin-6 (IL-6) receptor antibodies (e.g., tocilizumab or sarilumab)^11,12^, and Janus kinase (JAK) inhibitors (e.g., baricitinib)^13,14^, the incidence of severe ARDS and related mortality remain a major concern in hospitalized COVID-19 patients.

The principal therapy for hospitalized patients with moderate to severe ARDS is supportive care and high-flow oxygen administration progressing to mechanical ventilation and subsequent use of extracorporeal membrane oxygenation (ECMO) in resilient cases. Hemodynamic support is employed for the management of septic shock^15^. However, these interventions are exclusively supportive, are usually administered when ARDS has advanced, and do not modify the physiological or immunological course of the disease. Consequently, ARDS remains a life-threatening condition associated with a high fatality rate.

Paridiprubart (developed under the name EB05) is a humanized immunoglobulin gamma (IgG) 1 kappa (κ) monoclonal antibody, which inhibits TLR4-mediated inflammatory processes. In a randomized, double-blind, placebo-controlled, Phase 1 study, healthy volunteers were given a single intravenous infusion of paridiprubart. The study demonstrated a good preliminary safety and tolerability profile for paridiprubart with no safety signal identified after administration of a dose of up to 15 mg/kg. Furthermore, in the in vivo LPS challenge portion of the study an infusion of 0.25 mg/kg paridiprubart was shown to block cytokine release (TNFα, IL-6, CXCL10 and IFNβ) and physiological responses (increased body temperature and heart rate) for up to 22 days post EB05 administration. To evaluate the efficacy and safety of paridiprubart, we conducted a multicenter, randomized, double-blind, placebo-controlled, Phase 2 trial in COVID-19 patients already receiving standard-of-care. Here, we report the results of a prespecified sub-study of paridiprubart for the treatment of very severe COVID-19 patients with ARDS receiving invasive mechanical ventilation and additional organ support such as pressors, renal replacement therapy (RRT), and/or ECMO.

## METHODS

### Trial Design

A multicenter, randomized, double-blind, placebo-controlled, Phase 2 study was performed in 44 hospitals across the USA, Canada, and Colombia to evaluate the safety and efficacy of paridiprubart in adult patients hospitalized with COVID-19. The study was comprised of: (1) a main study that was conducted on patients that were hospitalized with mild to severe COVID-19 disease defined as levels 3-6 on the 9-point WHO COVID-19 Severity Scale (WCSS) at baseline; and (2) a sub-study that was conducted on patients that had very severe COVID-19 disease defined as WCSS level 7 at baseline.

Here, we report the results of the sub-study carried out in 8 hospitals from the USA and Canada, which recruited severe COVID-19 patients receiving invasive mechanical ventilation and additional organ support such as pressors, renal replacement therapy (RRT), and/or extracorporeal membrane oxygenation (ECMO) between January 23, 2021, and August 5, 2021. The sub-study was approved by regulatory agencies from each jurisdiction and institutional review boards or ethics committees as part of the broader full Phase 2 trial. The study was conducted in accordance with Good Clinical Practice guidelines and Declaration of Helsinki.

### Data Safety Monitoring Board Oversight

The trial was overseen by an independent data and safety monitoring board (DSMB), with meetings planned for reviewing interim analyses of the sub-study data when 10 and then 20 deaths had occurred. The results of the first interim analysis showed no safety concern and a clinically important between group difference with respect to mortality rates, the primary endpoint of the sub-study. Consequently, the DSMB recommended that enrollment in the sub-study should be terminated for efficacy. The unblinded results showed the efficacy signal to be in favour of paridiprubart and the DSMB recommended continuation into a Phase 3 confirmatory trial.

### Patients

Eligible patients for the sub-study were 18 years old or older and had a confirmed SARS-CoV-2 infection with a WCSS of 7, meaning they were hospitalized with ARDS and required mechanical ventilation with additional organ support such as pressors, RRT or ECMO. Key exclusion criteria included pregnancy, active participation in other immunomodulator or immunosuppressant drug clinical trials, treatment with immunomodulator or immunosuppressant drugs which were not part of standard-of-care, and death being imminent/inevitable within the next 72 hours, irrespective of the provision of treatment. Written informed consent was required from all patients or their legal representative prior to performing any study-related screening assessments.

### Randomization and Blinding

Patients were randomized at a ratio of 1:1 to receive an infusion of either paridiprubart or placebo. All patients also received standard-of-care treatment per routine care at each participating site. Since standard-of-care for the management of COVID-19 differed across investigational sites, randomization was stratified by clinical site.

This study was double-blinded. At all times, treatment and randomization information was kept confidential and was not released to the investigator, the study staff, the contract research organization (JSS Medical Research Inc.), or the sponsor’s study team until after the conclusion of the study. The investigational product kits were blinded with an Interactive Web Response (IWR) system when shipped from the Drug Depot.

### Procedure

While in the ICU, patients received a single intravenous infusion of paridiprubart (15 mg/kg up to a maximum of 1440 mg) or a matching placebo over a period of 3 hours at baseline. Paridiprubart and matching placebo vials were manufactured and tested by Nova Laboratories Limited, Leicester, United Kingdom. All participants received standard-of-care treatment, which consisted of intensive care therapy in keeping with local clinical practice for COVID-19 and ARDS management and included concomitant medications such as corticosteroids, antivirals, and other treatments (such as vasopressors or anti-inflammatory drugs), lung protective ventilation, thrombosis prophylaxis, RRT, and ECMO, as required.

Baseline measurements were ascertained at the last assessment recorded at/or before study drug administration on Day 1. Follow-up was up to 60 days post-treatment with the investigational product. All assessments were carried out in-hospital until discharge. If the patient was discharged from the hospital before Day 28 or Day 60, all end of study assessments were done at the discharge visit and additional assessments were conducted by telephone.

The schedule of assessments is provided in the Supplementary Appendix.

### Outcomes

Efficacy endpoints of the sub-study included all-cause mortality at Days 28 and 60 post-treatment, survival time to Day 60, achievement of WCSS ≤3 at Day 28, improvement of ≥2 on the WCSS at Day 28, and ventilator-free days (VFD) at Day 28. Change in WCSS by Day 28 post-treatment was also added as an outcome for the purposes of this analysis.

Safety outcomes of the sub-study included the incidence of treatment-emergent adverse events (TEAEs), as well as their characterization in terms of seriousness and relationship to the study product.

### Statistical Methodology

Enrolment for the sub-study was to be completed when a total of 30 deaths had been observed. Based on the estimates available at the time of study design, approximately 100 patients were anticipated to be enrolled in the sub-study. The sample size calculation was based on the Cox proportional hazard for mortality at Day 60. The estimated cumulative mortality rate for the placebo and paridiprubart-treated groups was assumed to be 40% and 20%, respectively. The target hazard ratio for placebo/paridiprubart was 2.2892. Based on these assumptions, a total of 30 events (in approximately 100 patients) were required to be observed for 80% power to detect a statistically important effect defined as a one-sided alpha of 0.10 due to the exploratory nature of the proof of concept, Phase II sub-study.

Two interim analyses were planned for the study after 10 deaths and after 20 deaths. The Independent DSMB recommended termination of the sub-study after the first interim analysis due to the observed differences in mortality rates and distribution of total deaths between the two groups. Consequently, the study was terminated when a total of 33 patients had completed the study and 12 deaths were observed of which 9 were in the 28-day time frame.

Efficacy and safety were analyzed in the intent-to-treat population, defined as all randomized patients that received the study treatment. For participants who discontinued study participation due to transfer to another hospital, all available information was used to inform the efficacy and safety analyses, including survival status and date of death when applicable. Adverse events were inclusive of the entire follow-up period. The number of patients taking any concomitant medication as well as each medication were reported for the two treatment groups. All concomitant medications were coded according to the generic drug names using the WHO Drug Classifications (Global version B3/C3 or later).

Descriptive statistics were reported for patient demographic and baseline characteristics. Baseline patient and disease characteristics were compared between treatment groups with the independent samples Student’s t-test (or the Mann-Whitney test if the normality assumption was violated) for continuous variables and the Chi-Square test for categorical variables.

Between group differences with respect to all-cause mortality rates by Day 28 and Day 60 after treatment initiation was assessed with the odds ratio (OR) derived from simple logistic regression with bootstrapping, when possible, for estimation of the standard error of the parameter. Time to death was described with the Kaplan Meier estimator of the survival function with log-rank tests to compare the survival of the two groups. Cox’s Proportional Hazards models were used to produce estimates of the Hazard Ratios at 28- and 60-days. *Post hoc* subgroup analyses of all-cause mortality rate at Day 28, was stratified by (1) baseline Berlin ARDS severity, (2) baseline SOFA score, and (3) on-study corticosteroid use.

Between group differences with respect to the proportion achieving change in WCSS ≤ 3 and ≥ 2 points improvement in WCSS at Day 28 were assessed with the OR derived from simple logistic regression. For these endpoints missing data were imputed using a modified last-observation-carried-forward (LOCF) methodology, wherein: (1) Patients that died at any time during the study were classified as not having achieved the endpoint; (2) patients that were discharged early (prior to Day 28) and alive were classified as having reached the endpoint; (3) patients that were transferred to another institution had the last non-missing post baseline values used to determined achievement of the endpoint.

Between group differences with respect to the change in WCSS and VFD were assessed with the Independent Student’s t-test. For patients with missing 28-day WCSS the following imputations were used: (1) For patients that died, the 28-day WCSS was set as 8 (death); (2) patients that were discharged alive early (prior to Day 28) 28-day WCSS was set as 2 and (3) patients that were transferred to another institution the last non-missing post baseline values were used to replace the missing 28-day WCSS. VFDs were calculated as 28 – number of days on ventilation defined as last date on ventilation – first date on ventilation regardless of interruptions; for patients that died prior to day 28 VFD was set as 0.

TEAEs and serious TEAEs were coded according to MedDRA (version 24.1) and were reported by total number of events and system organ class.

## RESULTS

### Trial Population

A total of 33 patients with WCSS level 7 at baseline were enrolled in the sub-study of the trial across 8 North American hospitals. Of these patients, 13 were randomized to be treated with paridiprubart and 20 to receive placebo (Figure 1). All randomized patients received one dose of the study treatment and were included in the intent-to-treat (ITT) population analysis. One patient from the placebo group was lost to follow-up after hospital discharge prior to Day 28 and one patient from the paridiprubart group was lost to follow-up after hospital discharge prior to Day 60.

**Figure 1.**
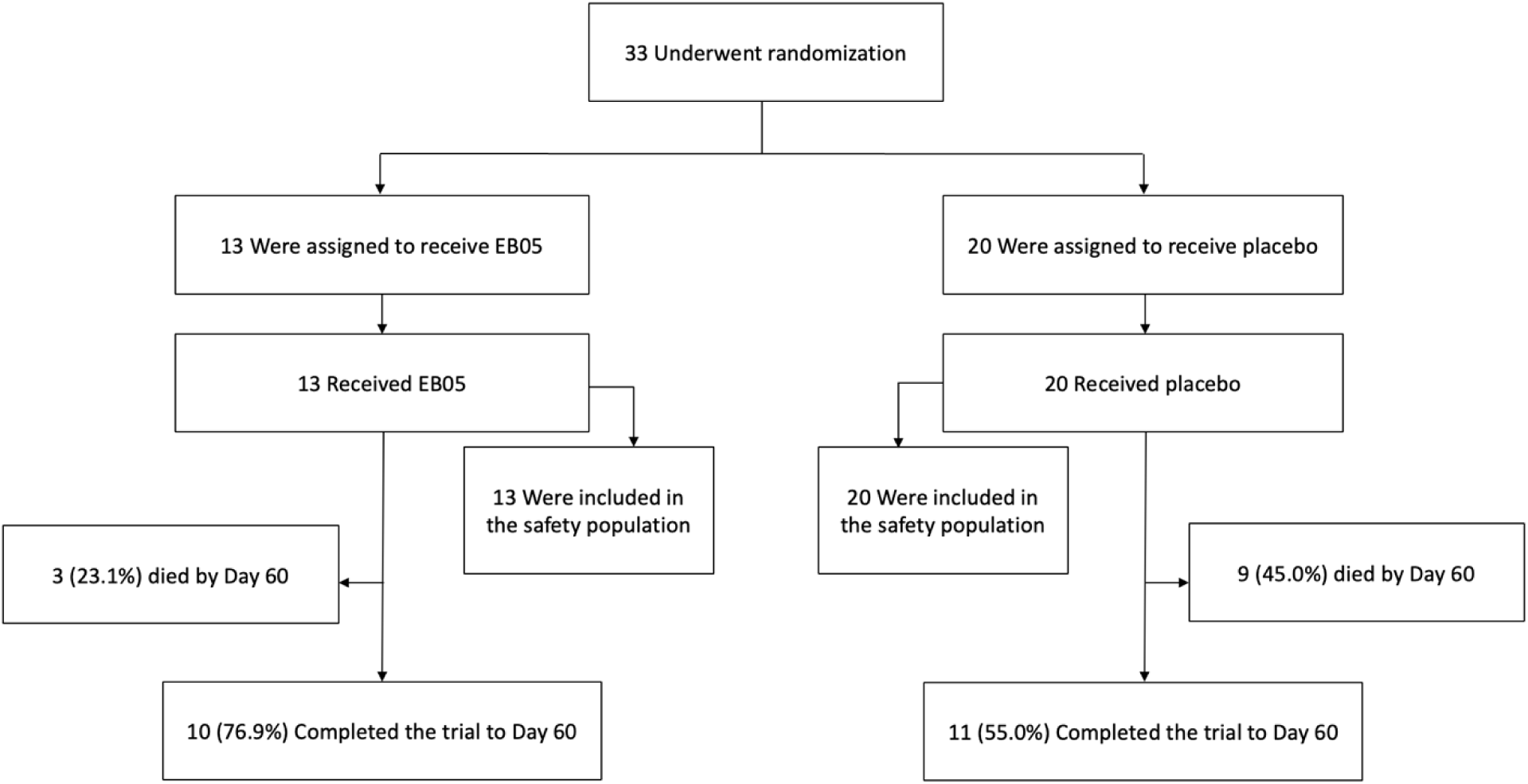
Patient Disposition. Results for all 33 randomized patients in the sub-study.

Baseline demographic and clinical characteristics were similar across the two groups of the sub-study (Table 1). The median age of patients was 48.0 years (interquartile range [IQR], 40.0 to 51.0) and 22 (66.7%) of the 33 patients were men. The majority (75.8%) of patients identified as white or Caucasian. Medical history of COPD and diabetes (without complications) was reported in 24.2% and 21.2% of patients, respectively. Paridiprubart-treated patients had slightly higher prevalence of acute kidney injury (AKI) at baseline (38.5% for paridiprubart vs. 20.0% for placebo). The proportion of patients with moderate to severe ARDS (100% for paridiprubart vs. 90% for placebo) and mean (SD) SOFA score at baseline: (9.9 ± 2.9 for paridiprubart vs. 9.6 ± 2.8 for placebo) were similar between the groups. A majority (81.8%) of patients were on ECMO at baseline. Concurrent standard-of-care treatments included corticosteroids (66.7% of patients), immune modulators (3.0%), and antivirals (6.1%). A higher proportion of patients in the paridiprubart group were treated by antivirals (15.4% vs. 0.0%; P=0.070) and immune modulators (7.7% vs. 0.0%), while a higher proportion of patients in the placebo group were treated with corticosteroids (75.0% vs. 53.8%; P=0.208)

**Table 1.** Demographic and Clinical Characteristics of the Patients at Baseline.

| Characteristic | Paridiprubart<br>(N = 13) | Placebo<br>(N = 20) | All Patients<br>(N = 33) | P value |
| --- | --- | --- | --- | --- |
| Age, years — median (IQR) | 46.0 (35.0-51.0) | 48.5 (40.5-51.5) | 48.0 (40.0-51.0) | 0.388<sup>^</sup> |
| Male sex — no. (%) | 9 (69.2) | 13 (65.0) | 22 (66.7) | 0.801* |
| Race or ethnic group — no. (%) |  |  |  | 0.349* |
| Asian | 4 (30.8) | 2 (10.0) | 6 (18.2) |  |
| Black or African American | 0 (0.0) | 1 (5.0) | 1 (3.0) |  |
| Not Specified/Unknown | 0 (0.0) | 1 (5.0) | 1 (3.0) |  |
| White or Caucasian | 9 (69.2) | 16 (80.0) | 25 (75.8) |  |
| Medical History — no. (%) |  |  |  |  |
| COPD | 2 (15.4) | 6 (30.0) | 8 (24.2) | 0.338* |
| Diabetes (without complications) | 2 (15.4) | 5 (25.0) | 7 (21.2) | 0.509* |
| Diabetes (with complications) | 2 (15.4) | 1 (5.0) | 3 (9.1) | 0.311* |
| Myocardial infarction | 1 (7.7) | 1 (5.0) | 2 (6.1) | 0.751* |
| Liver disease (moderate or severe) | 0 (0.0) | 1 (5.0) | 1 (3.0) | 0.413* |
| Peptic ulcer | 1 (7.7) | 0 (0.0) | 1 (3.0) | 0.208* |
| Renal disease | 0 (0.0) | 1 (5.0) | 1 (3.0) | 0.413* |
| WCCS level — no. (%) |  |  |  |  |
| 7: Ventilation + additional organ support (pressors, RRT, ECMO) | 13 (100.0) | 20 (100.0) | 33 (100.0) |  |
| AKIN stage — no. (%) |  |  |  | 0.063* |
| None | 8 (61.5) | 16 (80.0) | 24 (72.7) |  |
| Stage 1 | 4 (30.8) | 1 (5.0) | 5 (15.2) |  |
| Stage 2 | 1 (7.7) | 0 (0.0) | 1 (3.0) |  |
| Stage 3 | 0 (0.0) | 3 (15.0) | 3 (9.1) |  |
| Berlin ARDS severity — no. (%) |  |  |  | 0.500* |
| Mild | 0 (0.0) | 2 (10.0) | 2 (6.1) |  |
| Moderate | 3 (23.1) | 4 (20.0) | 7 (21.2) |  |
| Severe | 10 (76.9) | 14 (70.0) | 24 (72.7) |  |
| SOFA score — mean (SD) | 9.9 (2.9) | 9.6 (2.8) | 9.7 (2.8) | 0.807^ |
| Other treatments — no. (%) |  |  |  |  |
| ECMO | 12 (92.3) | 15 (75.0) | 27 (81.8) | 0.208* |
| Corticosteroids<sup>†</sup> | 7 (53.8) | 15 (75.0) | 22 (66.7) | 0.208* |
| Immune modulators<sup>†</sup> | 1(7.7) | 0 (0.0) | 1 (3.0) | 0.208* |
| Antivirals<sup>†</sup> | 2 (15.4) | 0 (0.0) | 2 (6.1) | 0.070* |

<sup>†</sup>: At baseline or during study.
^: P-value based on Student's t-test for Independent groups.
\*: P-value based on 2-sided Pearson Chi-Square.

### Survival

The patients treated with paridiprubart had significantly lower all-cause mortality rates by Day 28 post-treatment when compared to placebo, 7.7% (1/13) versus 40.0% (8/20), respectively (Odds Ratio [OR]=0.125; 95% confidence interval [CI], 0.013 to 1.160; P=0.06; P[bootstrap]=0.011). The 28-day mortality Risk Difference between paridiprubart and placebo was −32.3% (95% CI, −57.0 to −0.1; P=0.042) (Table 2). Lower 60-day mortality rate was also observed in the paridiprubart-treated patients when compared to placebo: 23.1% (3/13) versus 45% (9/20), respectively (OR=0.367; 95% CI, 0.077 to 1.749; P=0.208; P[bootstrap]=0.162). The 60-day mortality Risk Difference between paridiprubart and placebo was −21.9 (95% CI, −59.9 to 16.0; P=0.201) (Table 2).

**Table 2.**
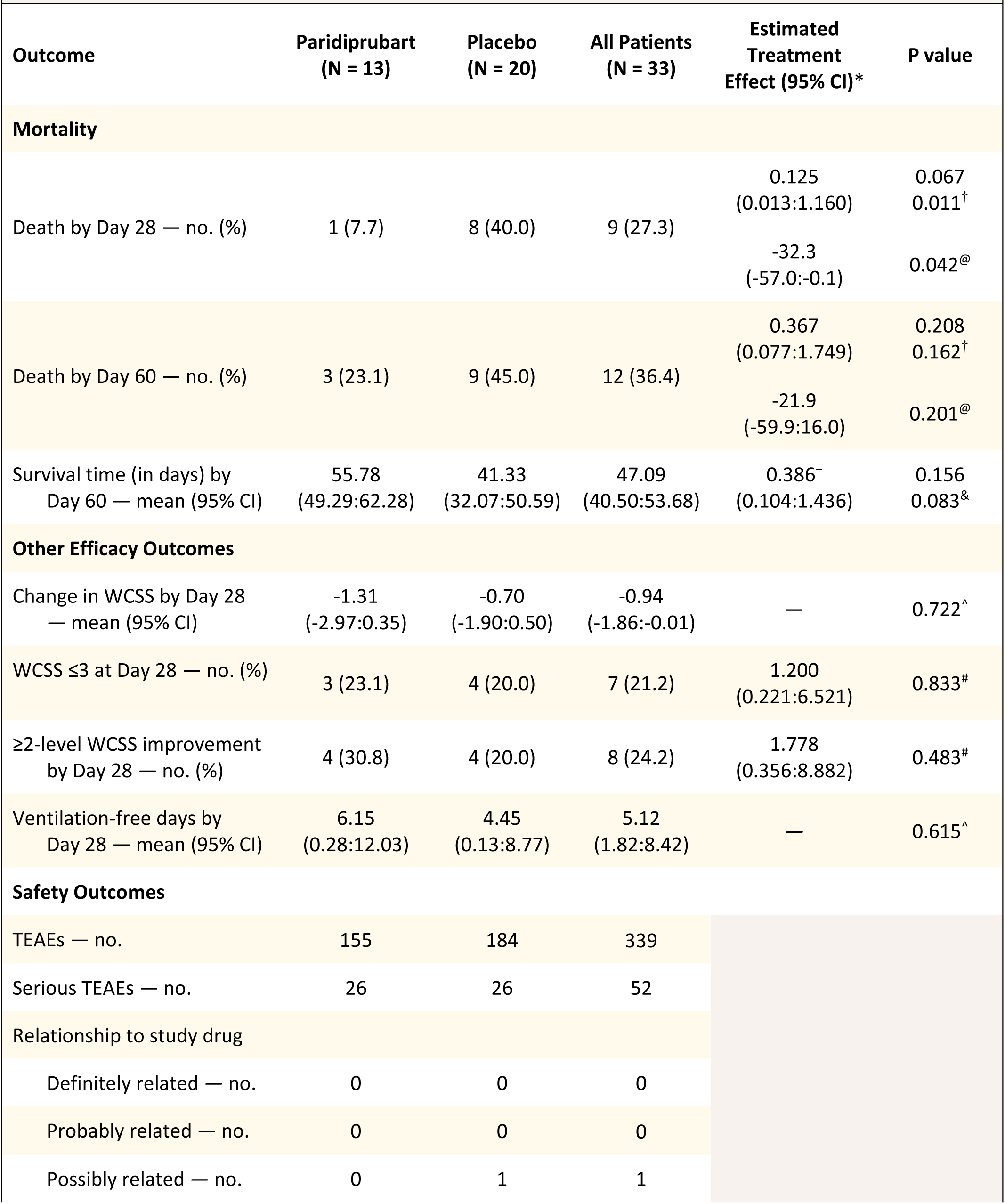
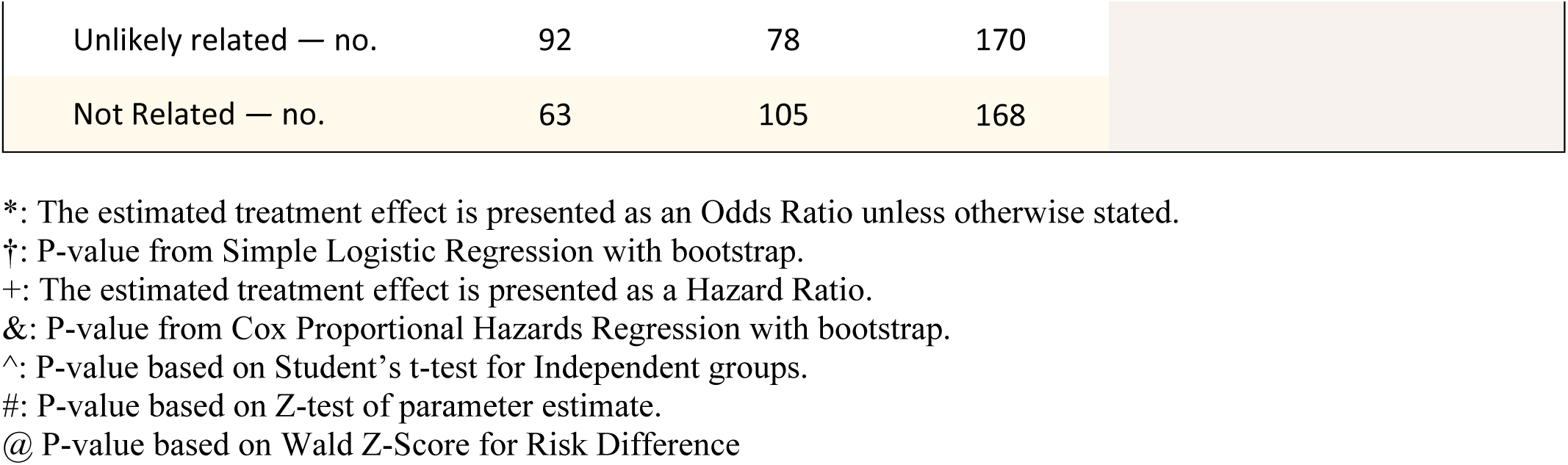
Efficacy and Safety Outcomes.

| Outcome | Paridiprubart<br>(N = 13) | Placebo<br>(N = 20) | All Patients<br>(N = 33) | Estimated<br>Treatment<br>Effect (95% CI)* | P value |
| --- | --- | --- | --- | --- | --- |
| Mortality |  |  |  |  |  |
| Death by Day 28 — no. (%) | 1 (7.7) | 8 (40.0) | 9 (27.3) | 0.125<br>(0.013:1.160) | 0.067<br>0.011<sup>†</sup> |
|  |  |  |  | -32.3<br>(-57.0:-0.1) | 0.042<sup>@</sup> |
| Death by Day 60 — no. (%) | 3 (23.1) | 9 (45.0) | 12 (36.4) | 0.367<br>(0.077:1.749) | 0.208<br>0.162<sup>†</sup> |
|  |  |  |  | -21.9<br>(-59.9:16.0) | 0.201<sup>@</sup> |
| Survival time (in days) by<br>Day 60 — mean (95% CI) | 55.78<br>(49.29:62.28) | 41.33<br>(32.07:50.59) | 47.09<br>(40.50:53.68) | 0.386<sup>+</sup><br>(0.104:1.436) | 0.156<br>0.083<sup>&</sup> |
| Other Efficacy Outcomes |  |  |  |  |  |
| Change in WCSS by Day 28<br>— mean (95% CI) | -1.31<br>(-2.97:0.35) | -0.70<br>(-1.90:0.50) | -0.94<br>(-1.86:-0.01) | — | 0.722<sup>^</sup> |
| WCSS ≤3 at Day 28 — no. (%) | 3 (23.1) | 4 (20.0) | 7 (21.2) | 1.200<br>(0.221:6.521) | 0.833<sup>#</sup> |
| ≥2-level WCSS improvement<br>by Day 28 — no. (%) | 4 (30.8) | 4 (20.0) | 8 (24.2) | 1.778<br>(0.356:8.882) | 0.483<sup>#</sup> |
| Ventilation-free days by<br>Day 28 — mean (95% CI) | 6.15<br>(0.28:12.03) | 4.45<br>(0.13:8.77) | 5.12<br>(1.82:8.42) | — | 0.615<sup>^</sup> |
| Safety Outcomes |  |  |  |  |  |
| TEAEs — no. | 155 | 184 | 339 |  |  |
| Serious TEAEs — no. | 26 | 26 | 52 |  |  |
| Relationship to study drug |  |  |  |  |  |
| Definitely related — no. | 0 | 0 | 0 |  |  |
| Probably related — no. | 0 | 0 | 0 |  |  |
| Possibly related — no. | 0 | 1 | 1 |  |  |

|  |  |  |  |
| --- | --- | --- | --- |
| Unlikely related — no. | 92 | 78 | 170 |
| Not Related — no. | 63 | 105 | 168 |

\*: The estimated treatment effect is presented as an Odds Ratio unless otherwise stated.
†: P-value from Simple Logistic Regression with bootstrap.
+: The estimated treatment effect is presented as a Hazard Ratio.
&: P-value from Cox Proportional Hazards Regression with bootstrap.
^: P-value based on Student's t-test for Independent groups.

The Kaplan Meier estimated overall mean survival time was higher for the paridiprubart-treated patients than for placebo, with 55.8 days (95% CI, 49.3 to 62.3) versus 41.3 days (95% CI, 32.1 to 50.6) (Table 2). Time to death results showed significantly higher 28-day survival for the paridiprubart-treated patients compared to placebo (Log-Rank P=0.0485) while the difference was statistically noteworthy for 60-day survival (Log-Rank P=0.137) (Figure 2). The 28-day and 60-day Hazard Ratios (HR) for paridiprubart/placebo were 0.163 (95% CI, 0.020 to 1.308; P=0.088; P[bootstrap]=0.054) and 0.386 (95% CI, 0.104 to 1.436; P=0.156; P[bootstrap]=0.083) (Table 2; Figure 2).

**Figure 2.**
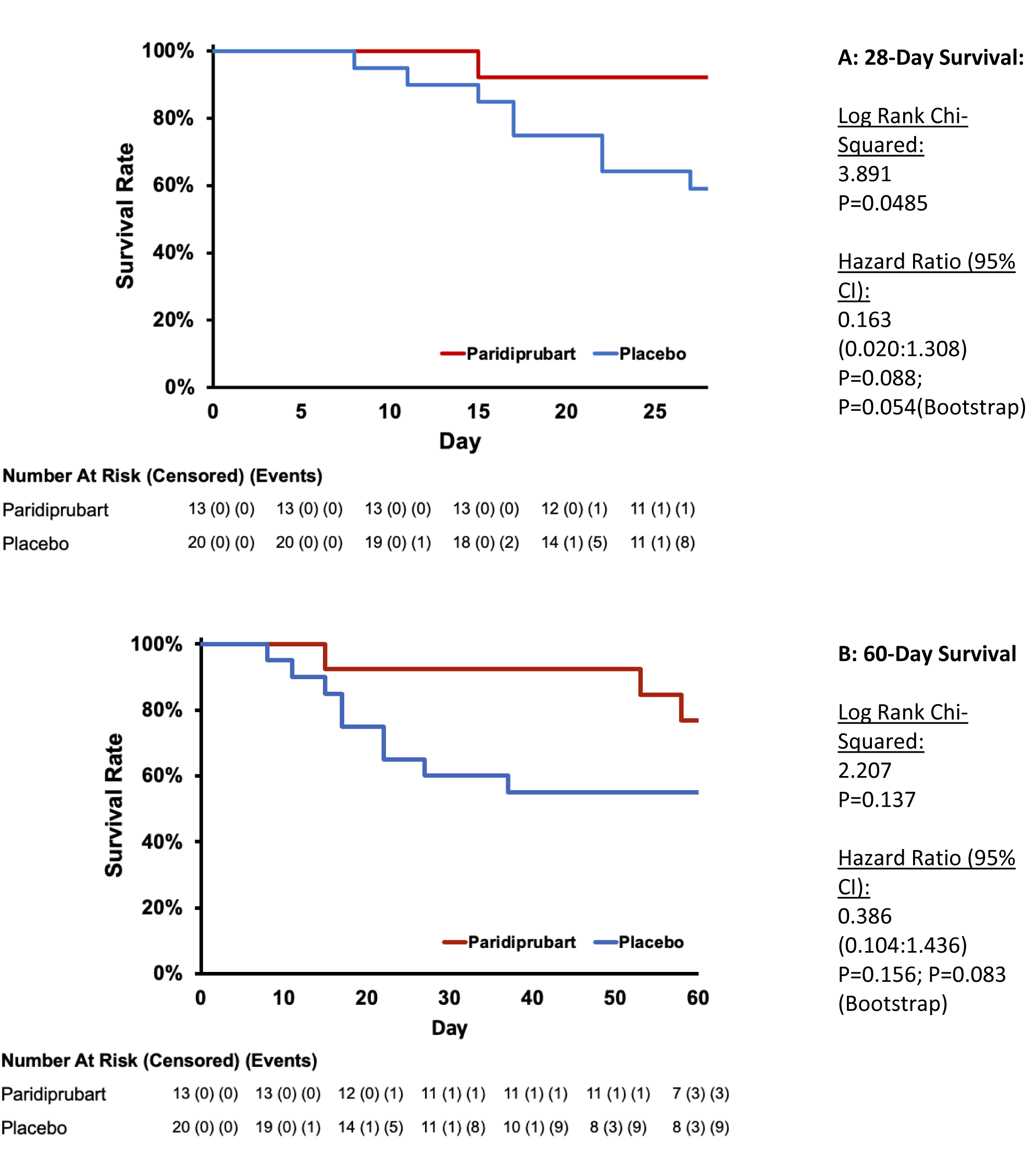
Kaplan Meier Survival Curves. Panel A: 28-day all-cause mortality; Panel B: 60-day all-cause mortality. Kaplan Meier estimator of the survival function with p-values based on log-rank tests for between group comparisons. Cox’s Proportional Hazards models were used to produce estimates of the Hazard Ratios.

### Other Efficacy Outcomes

Although not statistically significant (P=0.374) the decrease in WCSS was considerably higher in the paridiprubart group (−1.31; 95% CI, −2.97 to 0.35) when compared to the placebo group (−0.70; 95% CI, −1.90 to 0.50) (Table 2). The proportion of patients achieving WCSS ≤ 3 by Day 28 was similar for the two groups (paridiprubart: 23.1%; placebo: 20.0%; P=0.833) (Table 2). The proportion of patients achieving ≥2 grade decrease in WCSS by Day 28 was higher for paridiprubart when compared to placebo (30.8% vs. 20.0%; P=0.483) (Table 2). The mean ventilation-free days by Day 28 was higher for the paridiprubart-treated patients (6.15 days; 95% CI, 0.28 to 12.03) when compared to placebo (4.45 days; 95% CI, 0.13 to 8.77; P=0.607) (Table 2).

### Subgroup Analyses

Subgroup analyses of 28-day all-cause mortality showed lower risk for the paridiprubart-treated patients when compared to placebo across all strata. However, significantly lower 28-day mortality risk was observed in the paridiprubart group when compared to placebo for: patients with mild or moderate ARDS at baseline (Risk Difference=-0.52; 95% CI, −1.00 to −0.04; P=0.03); patients with baseline SOFA ≥ 10 (Risk Difference=-0.43; 95% CI, −0.79 to −0.08; P=0.02); and patients treated with corticosteroids (Risk Difference=-0.34; 95% CI, −0.64 to −0.05; P=0.02) (Figure 3).

**Figure 3.**
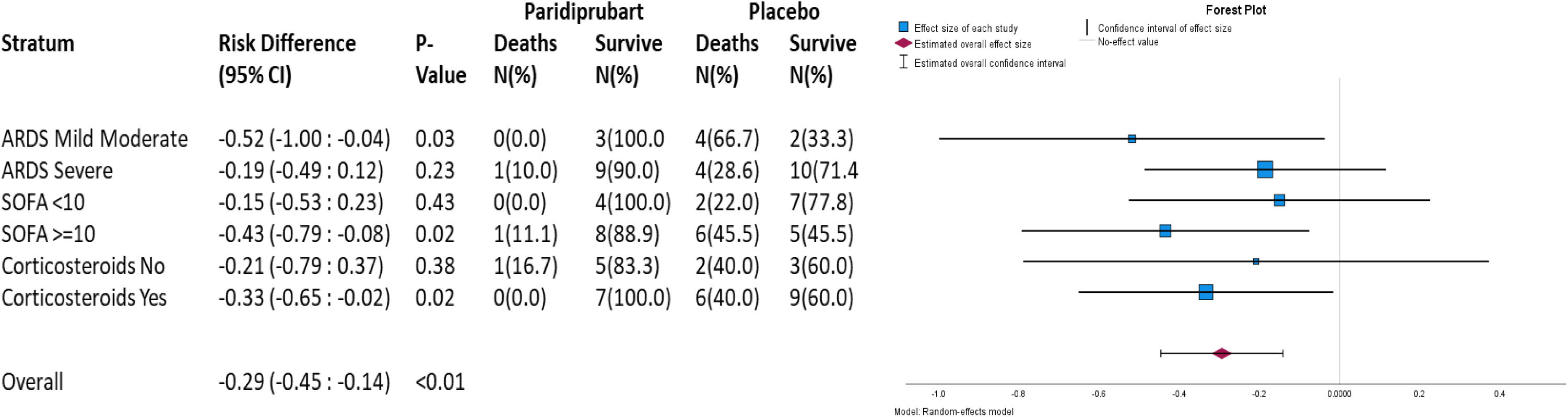
Subgroup Analyses for Mortality by Day 28. Mortality rate and risk difference (exact 95% confidence interval) for 28-day all-cause mortality for paridiprubart and placebo. P-values are based on Wald’s Z-score for independent proportions.

### Safety Outcomes

There was a total of 155 and 184 treatment emergent adverse events (TEAEs) in the paridiprubart and placebo groups, respectively, of which 26 in each treatment group were serious (Table 2; Table S1). All TEAEs in the study were categorized as “not related” or “unlikely related” to the study drug except for one, which was deemed “possibly related”, but occurred in the placebo group (Table 2).

## DISCUSSION

ARDS is a leading cause of mortality in critically ill patients with severe COVID-19. There is currently a paucity of treatments for ARDS, with therapeutic measures being mainly limited to supportive care, such as supplemental oxygen therapy, mechanical ventilation, and, as a last resort, ECMO. As such, there is a treatment gap with respect to the efficacious management of ARDS that can be administered earlier in the process with a disease modifying mechanism of action aimed at modulating the exaggerated immune response and cytokine storm. While ARDS is not exclusive to COVID-19^15–17^, its significant role in the causal pathway leading to death and the epidemiological acute burden presented by the pandemic has increased awareness and focus on the condition in general.

Paridiprubart has been shown to inhibit TLR4, a key transmembrane protein in the innate immune system, whose overactivation has been mechanistically implicated in the pathogenesis of ARDS. This Phase II study showed that a one-time infusion of paridiprubart is safe and efficacious in patients with severe COVID-19-related ARDS who were receiving invasive mechanical ventilation and/or additional organ support. One-time treatment with paridiprubart was associated with significantly reduced all-cause mortality and better clinical outcomes, with treatment benefit being consistently observed across several subgroups. There were no safety concerns related to treatment with paridiprubart.

Other than supportive care, treatments for severe COVID-19-related ARDS are currently limited to antivirals^18^, such as remdesivir^7,8^, or immune modulators, such as corticosteroids^9,10^, IL-6 antagonists^11,12^, or JAK inhibitors^13,14^; all of which have shown limited or mixed evidence for reducing all-cause mortality in clinical trials. Other therapies currently undergoing evaluation include mesenchymal stromal cell (MSC) therapy, which offers an option for multi-target immune modulation^19^. Initial smaller trials of MSC therapy in COVID-19-related ARDS patients provided supportive evidence of efficacy in reducing all-cause mortality^20–22^. However, these findings were not replicated in a recent randomized, double-blind, placebo-controlled, Phase 3 trial in 222 patients (30-day mortality: 37.5% [42/112] for MSC recipients versus 42.7% [47/110] for placebo recipients; relative risk, 0.88; 95% CI, 0.64 to 1.21; P=0.43)^23^, thereby, reinforcing the need for new therapeutics for this condition.

A main limitation of this sub-study was its small sample size. Positive interim efficacy data and lack of safety concern resulted in a recommendation from the DSMB to terminate enrolment in the Phase 2 trial prior to completion and to continue into a Phase 3 confirmatory trial. The reduced sample size of this study likely caused efficacy analyses to be underpowered. For instance, power calculations for the primary endpoint of all-cause mortality indicate that this endpoint was powered at 79% at 28-days post-treatment compared to only 39% at the 60-day time point. Despite the small sample size, the effects observed in this study were clinically important, with mortality-related endpoints achieving or approaching statistical significance. The strength of the effect was also supported by the distribution of total reported deaths between the two study groups (paridiprubart versus placebo) at each time point (1/9 versus 8/9 at Day 28; 3/12 versus 9/12 at Day 60). Other key strengths of this sub-study were its multicenter design, its homogeneous study population (i.e., WCSS=7), the selection of all-cause mortality as the primary outcome, and the consistency of the effect observed in favor of paridiprubart across endpoints and subgroups.

## CONCLUSION

Critically ill hospitalized COVID-19 patients with ARDS have limited treatment options. This Phase II trial sub-study demonstrated that paridiprubart may be efficacious in reducing all-cause mortality and improving clinical outcomes in this patient population. These results provide supporting evidence for a one-time infusion of paridiprubart in the treatment of COVID-19-induced ARDS. Although the current trial was aimed at assessing the management of patients with severe COVID-19-related ARDS, the results of the study have major implications in the management of ARDS in general, regardless of the causative factor or agent. A Phase 3 trial investigating the efficacy and safety of paridiprubart in a larger cohort of patients with COVID-19-induced ARDS is ongoing.

## Data Availability

All data produced in the present work are contained in the manuscript.

## ACKNOWLEDGEMENTS

The authors wish to acknowledge Vincent McCarty of JSS Medical Research for medical writing and editorial assistance during the preparation of this manuscript.

## EB05 Study Investigators

Ravindra Kashyap (Methodist Medical Center IL, Peoria IL, USA), George Diaz (Providence Regional Medical Center – Everett, Everett, WA, USA), J. W. Awori Hayanga (West Virginia University Medicine Heart & Vascular Institute, Morgantown, WV, USA), Fayed Mohamed (UCSF Fresno, San Francisco, CA, USA), Joseph Flynn (Norton Hospital, Louisville, KY,USA), Lorenzo Del Sorbo (Toronto General Hospital, Toronto, ON, Canada), Carlos Cervera (University of Alberta Hospital, Edmonton, AB, Canada), and Daniel Layish (AdventHealth Winter Park and AdventHealth Orlando, Orlando, FL, USA).

## Declarations of Competing Interests

B.G, M.B. and N.R. are employees of Edesa Biotech. All other authors declare no competing financial interests.

## Funding

Government of Canada’s Strategic Innovation Fund (SIF) and Edesa Biotech.

---

**Supplementary Table 1.**
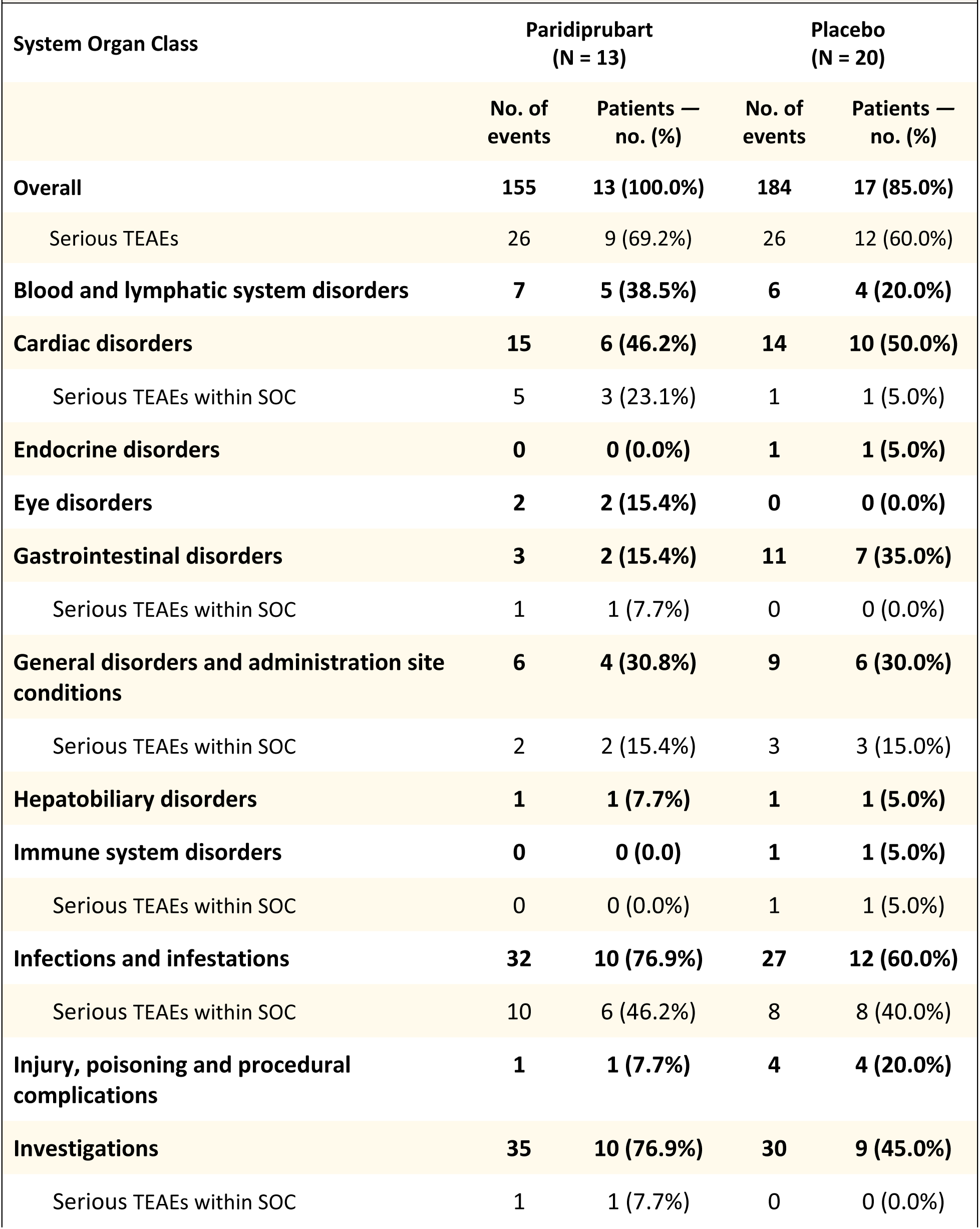
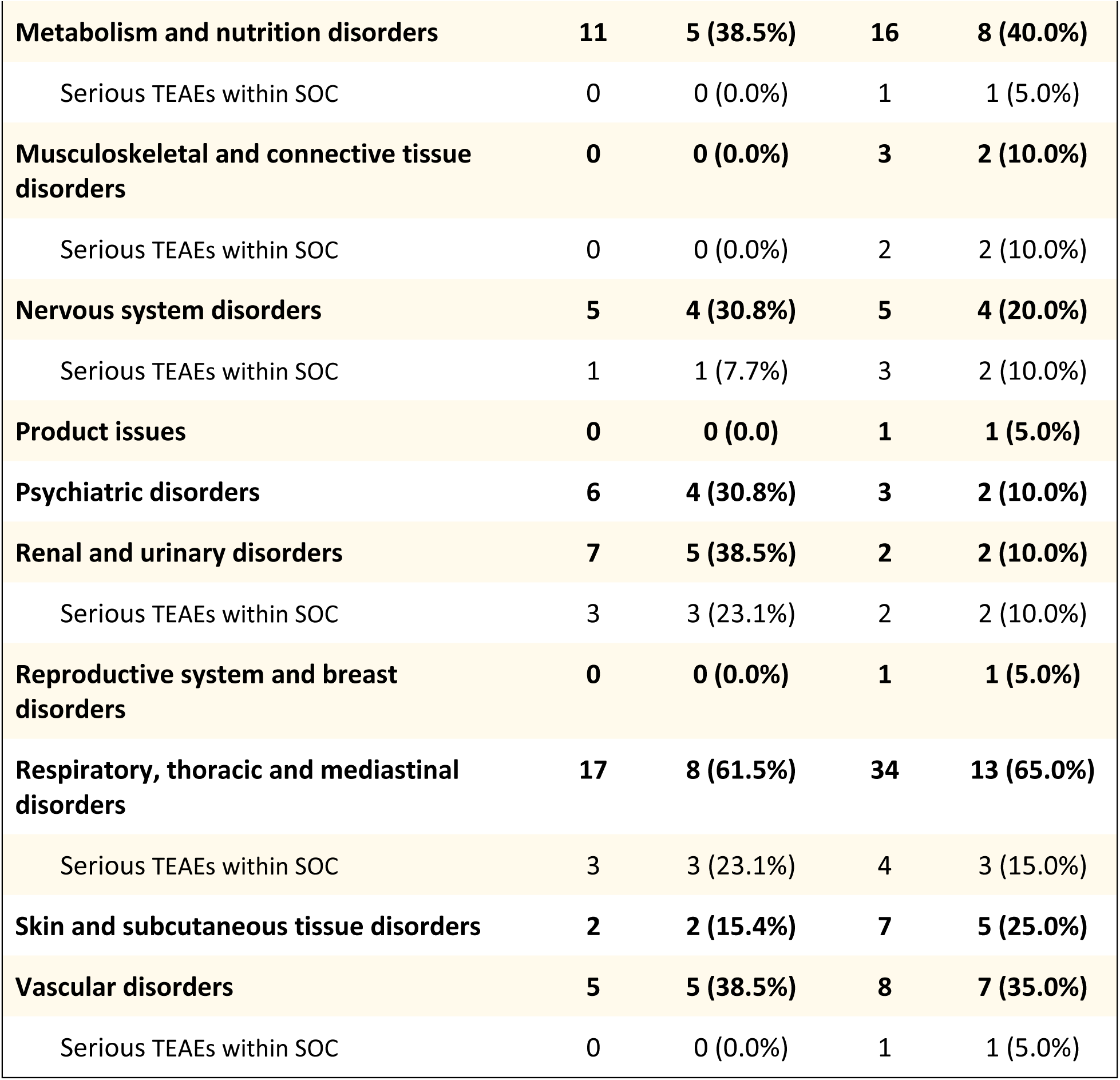
Incidence of Treatment-Emergent Adverse Events by System Organ Class and Treatment Group.

| System Organ Class | Paridiprubart<br>(N = 13) |  | Placebo<br>(N = 20) |  |
| --- | --- | --- | --- | --- |
|  | No. of<br>events | Patients —<br>no. (%) | No. of<br>events | Patients —<br>no. (%) |
| **Overall** | **155** | **13 (100.0%)** | **184** | **17 (85.0%)** |
| Serious TEAEs | 26 | 9 (69.2%) | 26 | 12 (60.0%) |
| **Blood and lymphatic system disorders** | **7** | **5 (38.5%)** | **6** | **4 (20.0%)** |
| **Cardiac disorders** | **15** | **6 (46.2%)** | **14** | **10 (50.0%)** |
| Serious TEAEs within SOC | 5 | 3 (23.1%) | 1 | 1 (5.0%) |
| **Endocrine disorders** | **0** | **0 (0.0%)** | **1** | **1 (5.0%)** |
| **Eye disorders** | **2** | **2 (15.4%)** | **0** | **0 (0.0%)** |
| **Gastrointestinal disorders** | **3** | **2 (15.4%)** | **11** | **7 (35.0%)** |
| Serious TEAEs within SOC | 1 | 1 (7.7%) | 0 | 0 (0.0%) |
| **General disorders and administration site conditions** | **6** | **4 (30.8%)** | **9** | **6 (30.0%)** |
| Serious TEAEs within SOC | 2 | 2 (15.4%) | 3 | 3 (15.0%) |
| **Hepatobiliary disorders** | **1** | **1 (7.7%)** | **1** | **1 (5.0%)** |
| **Immune system disorders** | **0** | **0 (0.0)** | **1** | **1 (5.0%)** |
| Serious TEAEs within SOC | 0 | 0 (0.0%) | 1 | 1 (5.0%) |
| **Infections and infestations** | **32** | **10 (76.9%)** | **27** | **12 (60.0%)** |
| Serious TEAEs within SOC | 10 | 6 (46.2%) | 8 | 8 (40.0%) |
| **Injury, poisoning and procedural complications** | **1** | **1 (7.7%)** | **4** | **4 (20.0%)** |
| **Investigations** | **35** | **10 (76.9%)** | **30** | **9 (45.0%)** |
| Serious TEAEs within SOC | 1 | 1 (7.7%) | 0 | 0 (0.0%) |
| **Metabolism and nutrition disorders** | **11** | **5 (38.5%)** | **16** | **8 (40.0%)** |
| Serious TEAEs within SOC | 0 | 0 (0.0%) | 1 | 1 (5.0%) |
| **Musculoskeletal and connective tissue disorders** | **0** | **0 (0.0%)** | **3** | **2 (10.0%)** |
| Serious TEAEs within SOC | 0 | 0 (0.0%) | 2 | 2 (10.0%) |
| **Nervous system disorders** | **5** | **4 (30.8%)** | **5** | **4 (20.0%)** |
| Serious TEAEs within SOC | 1 | 1 (7.7%) | 3 | 2 (10.0%) |
| **Product issues** | **0** | **0 (0.0)** | **1** | **1 (5.0%)** |
| **Psychiatric disorders** | **6** | **4 (30.8%)** | **3** | **2 (10.0%)** |
| **Renal and urinary disorders** | **7** | **5 (38.5%)** | **2** | **2 (10.0%)** |
| Serious TEAEs within SOC | 3 | 3 (23.1%) | 2 | 2 (10.0%) |
| **Reproductive system and breast disorders** | **0** | **0 (0.0%)** | **1** | **1 (5.0%)** |
| **Respiratory, thoracic and mediastinal disorders** | **17** | **8 (61.5%)** | **34** | **13 (65.0%)** |
| Serious TEAEs within SOC | 3 | 3 (23.1%) | 4 | 3 (15.0%) |
| **Skin and subcutaneous tissue disorders** | **2** | **2 (15.4%)** | **7** | **5 (25.0%)** |
| **Vascular disorders** | **5** | **5 (38.5%)** | **8** | **7 (35.0%)** |
| Serious TEAEs within SOC | 0 | 0 (0.0%) | 1 | 1 (5.0%) |

## Notes

### Competing Interest Statement

BG, MB, and NR are employees of
Edesa Biotech. All other authors declare no competing financial interests.

### Clinical Trial

NCT04401475

### Funding Statement

The Strategic Innovation Fund from the Government of Canada and Edesa Biotech.

### Author Declarations

Ethics committee and IRB of Methodist Medical Center IL, Providence Regional Medical Center, West Virginia University Medicine Heart and Vascular Institute, Morgantown, UCSF Fresno, Norton Hospital, Toronto General Hospital, University of Alberta Hospital, and AdventHealth Winter Park and AdventHealth Orlando gave ethical approval for this work.

